# Amygdala hyperactivation relates to eating behavior: a potential indicator of food addiction in Prader-Willi syndrome

**DOI:** 10.1101/2022.08.03.22278273

**Authors:** Kuzma Strelnikov, Jimmy Debladis, Juliette Salles, Marion Valette, Julie Cortadellas, Maithé Tauber, Pascal Barone

**Affiliations:** Brain & Cognition Research Center (CerCo), University of Toulouse Paul Sabatier, Toulouse, France; Brain & Cognition Research Center (CerCo), CNRS, Toulouse, France; ENT Department, Purpan Hospital, Toulouse, France; Department of Psychiatry, University Hospital of Toulouse, CHU Toulouse, France; Institut Toulousain des Maladies Infectieuses et Inflammatoires (Infinity) INSERM UMR1291 - CNRS UMR5051 - Université Toulouse III, Toulouse, France; Prader-Willi Syndrome Reference Center, Children’s Hospital-INSERM-University of Toulouse Paul Sabatier, Toulouse, France; UMR 1027 Inserm - Paul Sabatier University, Toulouse, France

**Keywords:** addiction, Prader-Willi syndrome, eating behavior, amygdala

## Abstract

Prader-Willi syndrome (PWS) is a rare genetic neurodevelopmental disorder involving nutritional, endocrine /metabolic, emotional and behavior dimensions. There is evidence for impaired hypothalamic development and function in PWS, involving oxytocin and ghrelin, which can account for the typical PWS phenotype. Hyperphagia with addiction-like behavior is one of the common features of PWS and is a consequence of the hypothalamic dysfunction. In this study, we hypothesized that brain regions associated with compulsive eating behavior would be abnormally activated by food-related odors in PWS, as these can stimulate the appetite and induce hunger-related behavior.

**Methods:** We used a classic olfactory discrimination test to verify that olfaction was normal in patients with PWS. In an fMRI scanner, we presented two odors, a tulip and a caramel odor, which have a different hedonic valence and a different capacity to arouse hunger-related behavior.

**Results:** There was a five-fold higher activation in the right amygdala for the caramel odor compared with the tulip odor in patients with PWS (n=14). No such hyperactivation was found in age-matched controls (n=11). Cluster analysis of clinical hyperphagia scores in patients with PWS revealed a link with the right amygdala hyperactivation.

**Conclusions:** Our study provides evidence for functional alteration of the right amygdala in PWS, which is part of the brain reward network involved in food addiction. This finding may relate to dysfunction of the ghrelin and oxytocin systems in PWS, as these are involved in addictive behavior, appetite, and olfactory bulb regulation.

## Introduction

Prader-Willi syndrome (PWS) is a rare genetic neurodevelopmental disorder that results from the loss of gene expression of paternally inherited maternally imprinted genes of chromosome 15 at position q11-q13. In more than 55% of cases, there is a microdeletion (DEL) of this chromosome region that is inherited from the father; in 40–45% of cases, there is maternal uniparental disomy (UPD); and in less than 5% of cases, there is an imprinting defect or translocation involving this region. Our team recently confirmed that the incidence of PWS is around one in 20,000 births (1). PWS is characterized by hormone deficiencies and cognitive impairments, with intellectual disabilities that are usually mild. The syndrome is associated with a range of psychiatric symptoms, including emotional dysregulation and social cognitive deficits. There can also be other comorbidities, such as sleep-related breathing disorders, scoliosis, gastrointestinal disorders, dysautonomia, including hypersomnia, narcolepsy, and catatonia.

PWS is characterized by a specific trajectory that involves nutritional, endocrine/metabolic, and emotional/behavioral dimensions. There is a paradoxical sequence that starts with poor feeding with failure to thrive and anorexia, followed by unexplained, excessive weight gain, and then hyperphagia which leads to severe obesity (2, 3). In adulthood, there is compulsive food searching, storing, and hoarding. The constant drive for food is a major source of stress and anxiety for patients, families, and caregivers, and can prevent socialization (4). The hyperphagia could therefore be described as an addiction to food (5). A PWS-specific hyperphagia questionnaire (HQ) has been developed (6, 7) which assesses three relevant subscores/dimensions: i)behavior, relating to the times, frequency, actions, and bargaining to obtain food; ii) drive, relating to the strong desire (impulse) to speak about and consume food, and difficulty in diverting attention and avoiding frustration/anger; and iii) severity, relating to the extent to which food-related thoughts, words, and actions invade daily life.

The PWS phenotype has been shown to relate to impaired hypothalamic development and function (3). The hyperphagia may relate to abnormalities in oxytocin (OXT) and ghrelin, which are both linked to the hypothalamus. These hormones interact very closely and are involved in brain development and in regulating the dopaminergic reward system. OXT is secreted by hypothalamic neurons in the paraventricular and supraoptic nuclei, while ghrelin exerts most of its metabolic and appetite-regulating effects in the hypothalamus. In people with PWS as a group, the total plasma ghrelin levels are elevated from birth and throughout life (8). There are two forms of ghrelin: acylated (AG; fatty acid bound to the peptide chain) and un-acylated (UAG). AG has a strong orexigenic effect and is therefore called the “hunger hormone”, whereas UAG inhibits the effects of AG and has a resulting anorexigenic effect (9). It is possible that the hyperghrelinemia in PWS may play a role in the obsessive food-related thoughts and addiction to food, as well as in increasing patients’ appetites. The OXT system is also dysfunctional in patients with PWS. This hormone is known to be involved in emotion processing, and OXT administration has been found to increase trust in others and decrease sadness tendencies in patient with PWS (10, 11). It is possible that this hormone also plays a role in the hyperphagia in PWS.

Addictive behaviors involve different processes, such as incentive salience, stimulus-response habits, attentional bias, craving, executive functioning, and place- and emotional-conditioning (12). The sensory systems play an important role in these, especially in the case of incentive salience. Indeed, it has been found that the activation of sensory brain regions in response to drug-associated cues can predict relapse (13) and also correlates with craving, the severity of dependence, and automatized motor responses to the cues (14). The olfactory system plays an important role in nutrition and social behavior, and there is evidence that it is linked to the endocrine regulation of energy balance (15). Several studies have demonstrated that odors trigger an appetite for the relevant food (16-19), a phenomenon known as sensory-specific appetite. In this way, odors can alert us to food in our environment and orient our appetite accordingly (20, 21). There is evidence that the olfactory system may play a role in eating disorders. For instance, it has been found that overweight children fail to regulate food intake after exposure to an intense smell of tasty food (22), and patients with bulimia nervosa report a greater urge to binge eat after exposure to smell and taste cues (23).

In this study, we investigated the effect of food-related or unrelated odors on the activity of brain regions involved in reward (see (24) for review) in patients with PWS. We hypothesized that brain regions associated with compulsive eating behavior, namely the basal ganglia, extended amygdala, and prefrontal cortex, would be differentially activated in participants with PWS and controls.

## Methods and Materials

This clinical study (NCT0280437-33) was designed for adults with PWS and included brain imaging, neuropsychological assessments, the Hyperphagia Questionnaire (HQ), and a HQ that was modified to be used for clinical trials (HQ-CT). The study was approved by the local ethics committee (“Comité de Protection des Personnes Sud-Ouest et Outremer 1”, Toulouse Hospital CHU 13687203; National EudraCT 201300437-33). Written informed consent was obtained from each patient’s legal guardian or next of kin prior to the study. All of the patients participated in the study during routine follow-up visits to our center in Toulouse. Height, weight, body mass index (BMI), evaluation of eating behavior using the HQ, olfactory testing, and fMRI were all carried out in a one-day visit. Genetic subtypes were retrieved from the patients’ medical files. Sex- and age-matched controls were recruited for the study via a public announcement.

### Olfactory test

A standardized clinical test (25) was administered to evaluate the ability to recognize different odors. This involved presenting participants with sticks with different odors and asking them to complete a four-alternative forced-choice task (a card with four names of odors). Twelve different odor sticks were used; each was presented once.

### Visual detection task

A visual detection task was administered, as food-related behavior may be influenced by visual information as well as olfactory stimulation. For this task, pictures were used from the International Affective Picture System (26). The first set of pictures was of human faces; the second set included everyday objects and food; the third set contained the same images as the second set, but a Fourier transformation was applied to randomize the spectral content. There were a total of 12 images in each set, and these were presented in a random order. Participants were asked to respond as quickly as possible when they saw a picture by pressing a response key. Images were presented until the participants had responded. The stimuli were presented and responses were recorded using E-Prime software.

### fMRI procedure

The participants wore a mask and were asked to focus on odors that were sent through the mask. Two different olfactory stimuli were used: an odor related to food and an odor unrelated to food. For the former, we chose a caramel odor, because individuals with PWS have a preference for sweet foods; for the latter, we chose a tulip odor, because these flowers are not associated with food. The two odors were presented randomly, and there was no auditory or visual cue to alert participants to the onset of an odor. Each odor was presented in blocks that lasted 24 seconds. Within a block, the odor was presented for 4 seconds, followed by clean ambient air for 2 seconds (a purge). This was repeated four times (see Figure 1). The clean air was used to prevent olfactory bulb habituation to the odors. Between the blocks, clean air was sent for 12 seconds, and this was used as the baseline in the analysis.

**Figure 1.**
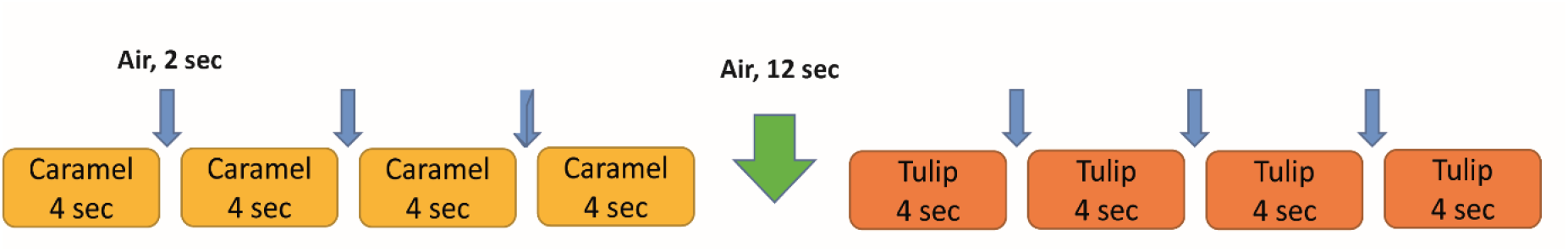
Experimental paradigm. The odors were presented for 4 seconds followed by clean air for 2 seconds (a purge). Between each block of odors, clean air was sent for 12 seconds; this was used as the baseline condition in the analysis.

In total, there were 56 blocks with the caramel odor, 48 blocks with the tulip odor, and 52 clean air baseline recordings. The total acquisition time was 16 minutes, which was divided into two 8-minute blocks with a small break in between.

The odors were presented using an olfactometer that was constructed based on a design from the De Volder team (29). This system has already been tested for use in MRI experiments (30). The system involves delivering odors through a 5-meter Teflon tube, which is directly connected to two vials that contain the odors. The olfactometer was connected to a computer, which controlled the opening and closing of valves. A scuba tank was used to continuously supply air and send the odors into the mask. The synchronization of the olfactory stimuli with the fMRI data was carried out using software developed by the De Volder team (29).

### Image acquisition parameters

BOLD and anatomical images were acquired using a 3T Philips ACHIEVA X-series MRI scanner. Anatomical images were obtained with T1-weighted gradient echo acquisition, a repetition time (TR) of 8.1 ms, an echo time (TE) of 3.7 ms, a flip angle of 8°, a field of view (FOV) of 240 × 240 mm, and a 1×1×1 mm resolution. For the BOLD signal images, T2* echo planar imaging (EPI) was used with a TR of 3 s, a TE of 30 ms, an acquisition time (TA) of 3 s, a flip angle of 90°, a FOV of 240 × 240 mm, a voxel size of 3×3×3 mm, and a matrix size of 80×80 pixels. Forty-five slices were acquired per volume with a total of 315 volumes; the first three volumes were discarded. The MRI session lasted about 30 minutes, 16 minutes of which were for the fMRI.

### fMRI acquisition and analysis

The MRI images were analyzed using SPM8 (www.fill.ion.ucl.ac.uk/spm/). The BOLD images were realigned for each subject, normalized to the MNI152 template, and smoothed using an isotropic Gaussian filter with a full width at half maximum (FWHM) of 6 mm.

We specified two regions of interests (ROI): the left and right amygdala, which are cytoarchitectonically defined. These ROIs were chosen because amygdala activity has been associated with the Yale Food Addiction Scale (27, 28), and also because amygdala activation has been observed for the anticipation of food (27). The ROIs were defined using the SPM Anatomy Toolbox.

We used k-means clustering (R stats library), an unsupervised machine learning algorithm, to partition the subjects into groups according to their hyperphagia scores. This was carried out because of the heterogeneous patient profiles and the large number of clinical scores. The k-means algorithm identifies a fixed number of clusters within a dataset, which aggregate together because of certain similarities in the scores. We predefined four clusters in order to approximate quartiles, which are widely used in the clinical literature. To stabilize the clustering results, 40 sets with random seeds were chosen for the machine learning. The resulting clusters (groups) of subjects were used to assess group effects for brain activity with ANOVA.

### Participants

A total of 16 participants with PWS were recruited for this study; one of these was excluded because of poor performance on the olfactory test and another was excluded because of technical problems in the MRI scanner. The analyses were therefore carried out for 14 patients, eight of whom had DEL and six of whom had UPD. The average age was 25±9 (SD) years, and there were seven women (Table1). Eleven controls were recruited for the study. The characteristics of the patients are shown in Table 1.

**Table 1.** Characteristics of the participants with PWS.

| **Patients** | **Genetic subtype** | **Sex** | **Age (years)** | **Total score HQ** | **Total HQ-CT** |
| --- | --- | --- | --- | --- | --- |
| 1 | UPD | F | 10-20 | 11 | 0 |
| 2 | DEL | F | 10-20 | 17 | 5 |
| 3 | UPD | F | 21-30 | 27 | 18 |
| 4 | DEL | M | 10-20 | 27 | 21 |
| 5 | DEL | F | 10-20 | 21 | 14 |
| 6 | DEL | F | 31-40 | 26 | 18 |
| 7 | DEL | M | 10-20 | 39 | 32 |
| 8 | UPD | M | 21-30 | 28 | 21 |
| 9 | DEL | F | 10-20 | 40 | 23 |
| 10 | UPD | M | 10-20 | 38 | 21 |
| 11 | UPD | M | 21-30 | 37 | 21 |
| 12 | DEL | M | 31-40 | 19 | 4 |
| 13 | DEL | M | 10-20 | 19 | 4 |
| 14 | UPD | F | 41-50 | 38 | 20 |

## Results

### Behavioral results

For the olfactory test, the average scores were 91±7.7% for the patients with PWS and 90±5.3% for the typically developed (TD) controls (see figure 2). All participants obtained scores above the normal threshold value of 80%. The scores did not differ significantly between two groups (t-test, p > 0.3).

**Figure 2.**
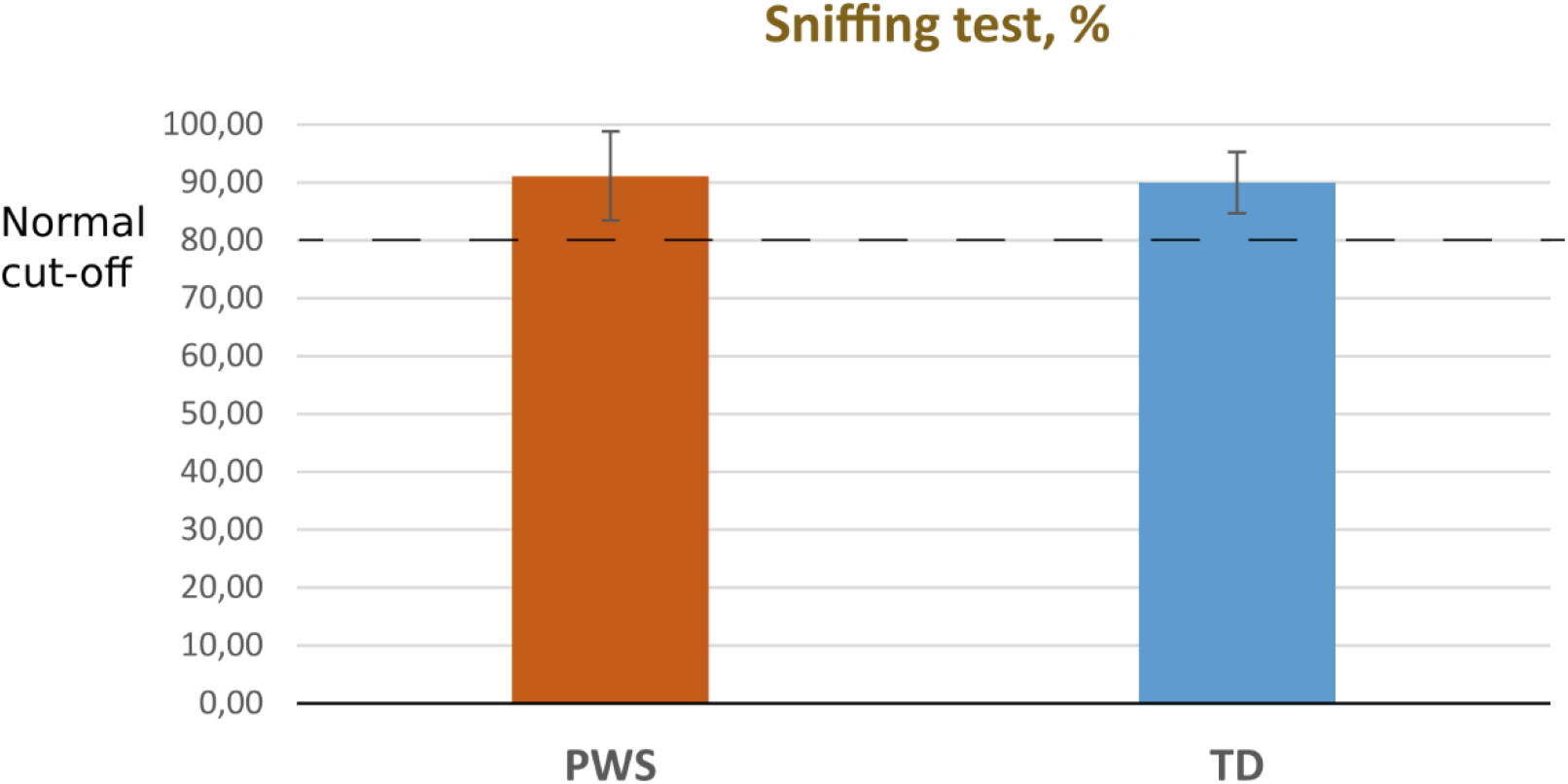
Olfactory test results for the PWS and TD control groups. All of the participants in the fMRI study achieved a score above the cut-off of 80%. There was no significant difference between the participants with PWS and the controls.

The visual test was run on the participants with PWS. The mean reaction time (RT) was 837±732 ms for the human face stimuli, 726±594 (SD) ms for the abstract images (with Fourier transformation), 895±757 ms for the everyday objects, and 731±737 ms for the food images. The differences were not statistically significant (bootstrap, p > 0.05). These results are in agreement with our previous study, where we found that there were no significant RT differences for different visual stimulus categories, including food, in a larger group of patients with PWS (31). In that study, it was also found that the subjects with PWS were significantly slower than TD controls, as is usual for any task.

### Group analysis of the neuroimaging data

To analyze brain responses to food-related olfactory stimulation, we began by assessing the effect of the caramel odor in a whole-brain analysis. For the participants with PWS, this approach revealed a single brain region that was significantly more activated by the caramel odor than by the tulip odor, which corresponded to the right amygdala (p_corr(FWE)_ < 0.05, (x,y,z) = (21,-1,-13); Figure 3A). To confirm the location of this activation, the region was compared with the anatomical position of the amygdala according to the cytoarchitectonic atlas (32), as illustrated in Figure 3B. Figure 3C shows the brain activation together with the anatomical location of the amygdala, revealing an overlap of around 40%.

**Figure 3.**
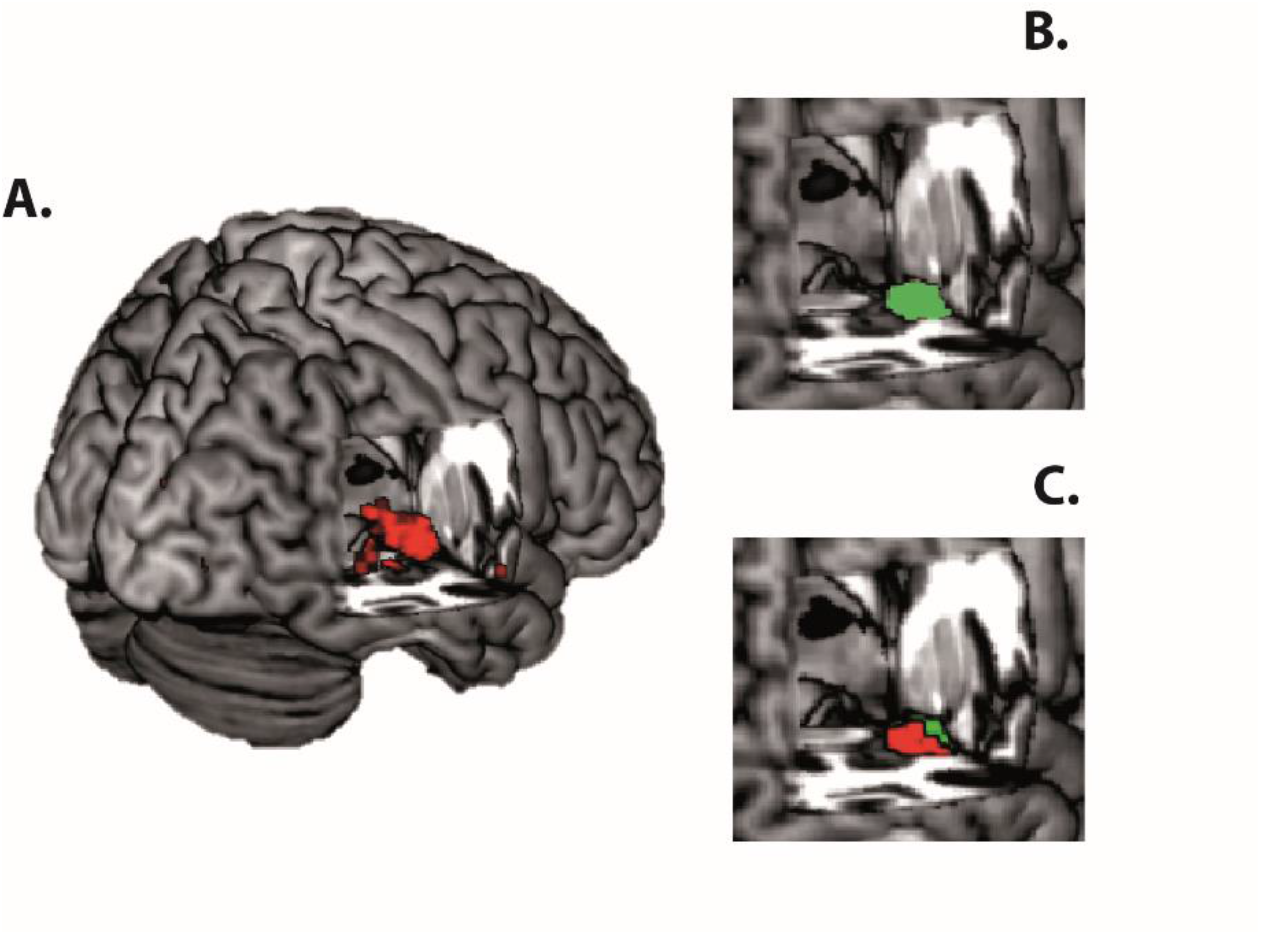
Specific BOLD activity in response to the caramel odor compared with the tulip odor. A. Difference between the caramel and tulip odor blocks in participants with PWS in the whole brain analysis. This pattern was significantly different compared with the control participants (pcorr < 0.05) B. Anatomical location of the right amygdala according to the cytoarchitectonic atlas. C. Overlap between the anatomical position of the right amygdala and the whole brain activity difference (in red).

Further analyses were carried out using a ROI centered on the amygdala, as described in the methods section. This approach was supported by the overlap described above, as well as by evidence that the amygdala plays a role in addictive behaviors (27). The analyses revealed a significant difference between the participants with PWS and the TD controls. Specifically, there was greater activation for the caramel odor compared with the tulip odor in the participants with PWS than in the controls (p_corr(FWE)_ < 0.05, (x,y,z)= (15, -1, -10)). We then extracted the peak activation values for each individual and compared the values for the caramel and tulip odors. Using a linear mixed effects model, we found that the participants with PWS had significantly greater activity in the amygdala region than the controls when presented with the caramel odor (p=0.02), but there was no significant group difference when they were presented with the tulip odor (p=0.59). In addition, there was a significant difference between the activity for the caramel and tulip odors in the PWS group (p=0.00056), but not in the TD controls (p=0.85; Figure 4A).

**Figure 4.**
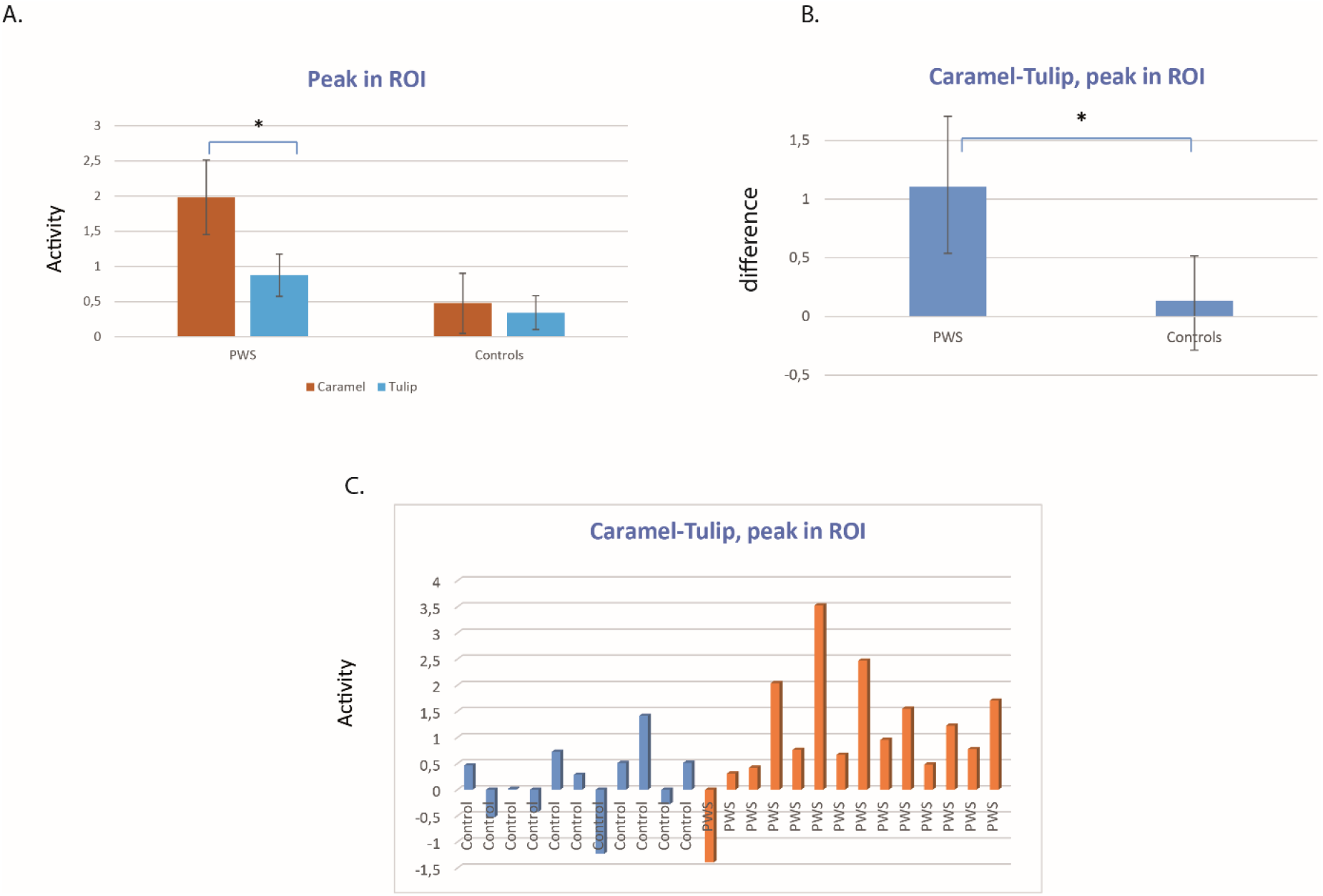
Activity in the right amygdala for the caramel and tulip odors. A. Peak activity in the right amygdala for the caramel and tulip odors in the PWS and control groups. B. Difference between the peak activity in the right amygdala for the caramel and tulip odors (“caramel – tulip”), shown for the PWS and control groups. C. Individual differences for the “caramel – tulip” right amygdala peak, shown for the participants with PWS and the controls.

Next, we subtracted the values for the tulip odor from those for the caramel odor, and compared the results for the two groups. The comparison revealed that the mean “caramel – tulip” activity difference was more than five times higher in the PWS group than in the controls (p < 0.05; Figure 4B). At an individual level, the “caramel – tulip” difference was found to be quite variable among the control participants, whereas a clear positive difference could be seen for the majority of the PWS participants, with only one subject manifesting a negative difference (Figure 4C). By defining difference values between -0.5 and 0.5 as statistical noise, eight of the 11 controls were seen to have no activity difference, whereas ten of the 14 PWS subjects had greater activity for the caramel odor and only one had greater activity for the tulip odor.

Interestingly, this latter patient also had the lowest HQ (total score 11) and HQ-CT (score 0) scores.

Analyses were carried out to determine whether there were differences for participants with the DEL or UPD genetic subtype of PWS. It was found that both subtype groups displayed a significant difference between the right amygdala activity peak for the caramel and tulip odors (Figure 5A). The “caramel – tulip” difference was not significantly different between the subtype groups (p > 0.05; Figure 5B), although the mean value was higher in the DEL group.

**Figure 5.**
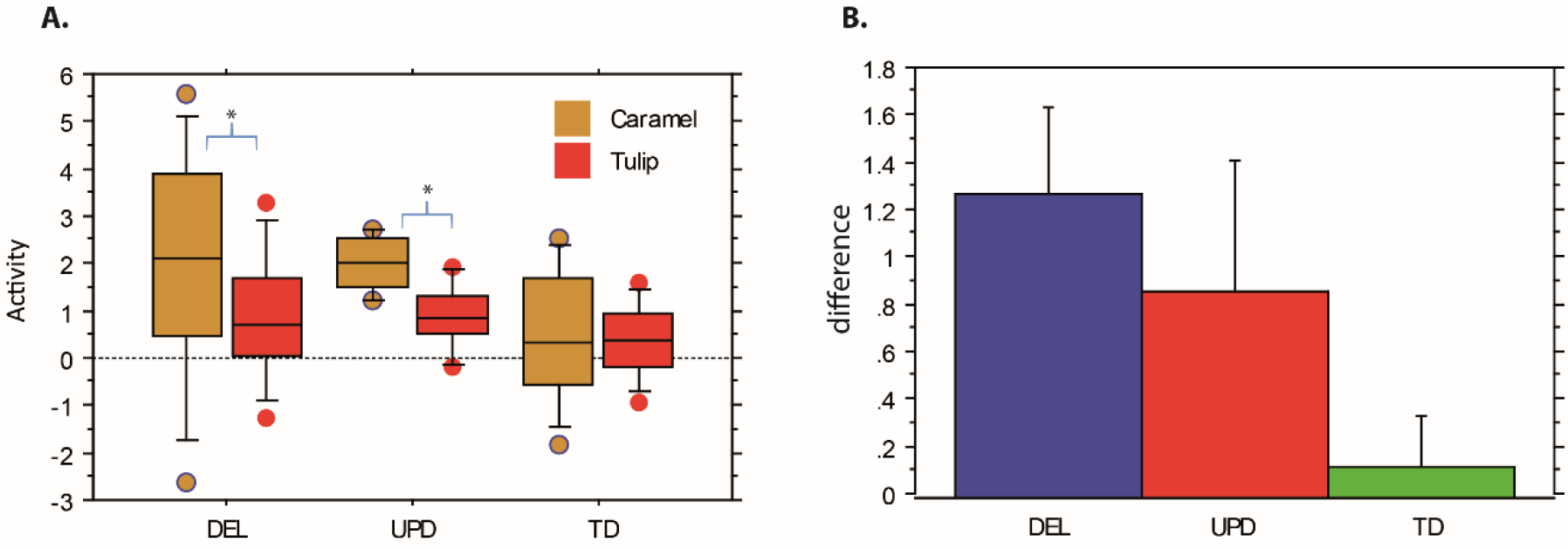
Right amygdala peak activity for participants with different genetic subtypes of PWS and TD controls. A. Right amygdala peak activity for the caramel and tulip odors in participants with the DEL or UPD subtypes of PWS and TD controls. No significant differences were found between the PWS subtype groups. B. Difference in amygdala activity in response to the caramel and tulip odors (“caramel – tulip”). There was no significant difference between the PWS subtype groups, although participants with the DEL subtype had the highest odor-related activity difference.

### Neuroimaging analysis for PWS clusters

We examined whether the individual differences in activity levels for the two different odors related to clinical scores of hyperphagia in the participants with PWS, specifically the total score and subscores of the HQ, and the total score of the HQ-CT. No significant correlations were found, probably because of the marked heterogeneity in the individual profiles. In order to further examine whether the clinical profiles related to the patterns of brain activity, we used k-means clustering to partition the subjects with PWS into groups according to their clinical scores. The machine learning algorithm detected three clusters with 3–5 subjects and one cluster with a single subject. The three main clusters were separated according to the first dimension (Figure 6A), which accounted for 84% of the variability in scores; the second dimension only accounted for 11% of the variability. There were strong correlations between the first dimension and both the total HQ score (r=0.995) and the HQ-CT score (r=0.986).

**Figure 6.**
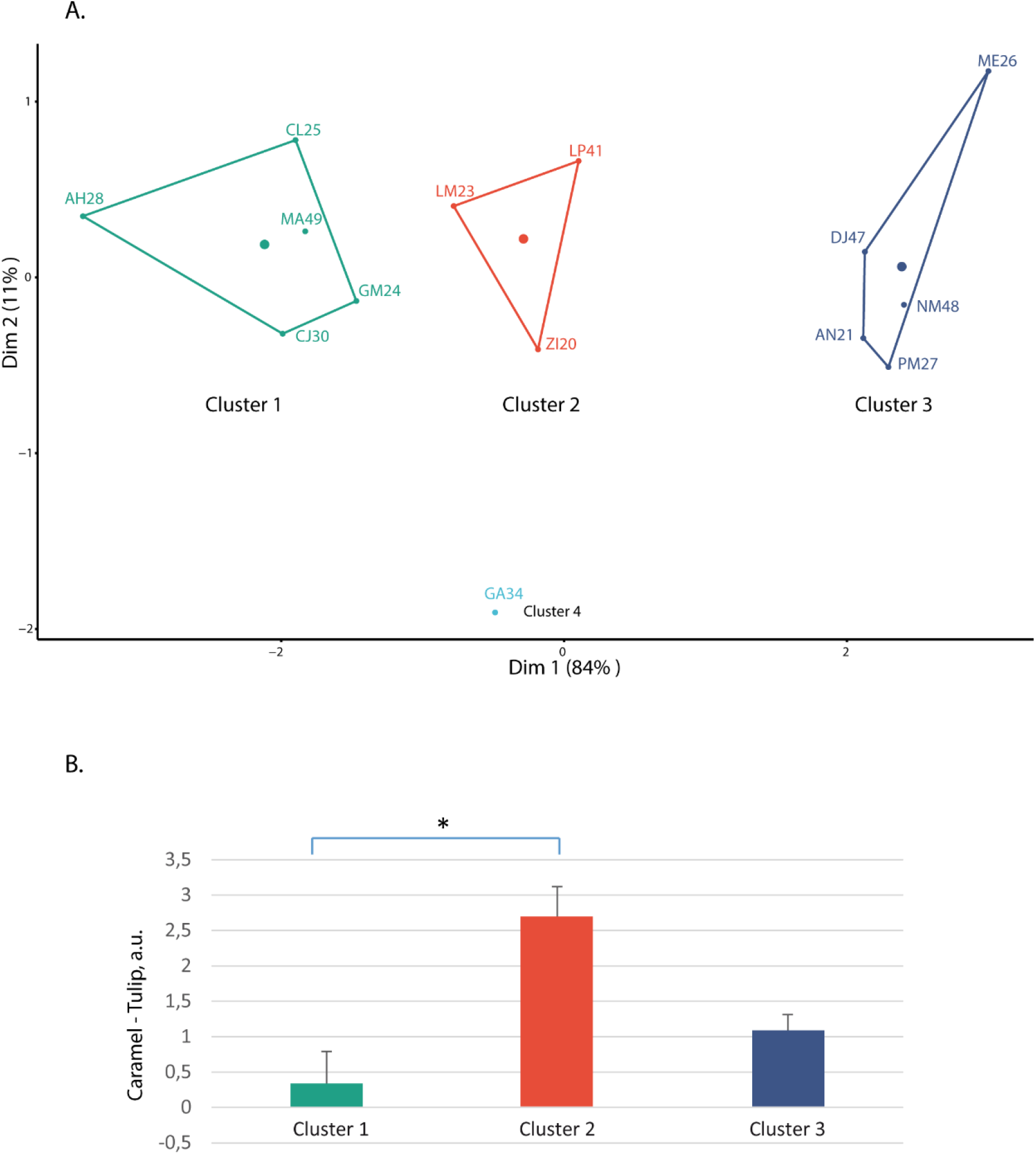
K-means clustering of scores and activity differences in the right amygdala. A. Unsupervised machine learning using k-means clustering detected four clusters of subjects according to their HQ and HQ-CT total scores. There were three main clusters with 3–5 subjects and a fourth cluster with one subject. The three main clusters were divided according to the first dimension, which accounted for 84% of the variability in scores. B. The “caramel - tulip” difference in right amygdala activity for the different clusters of subjects. There was a significant difference between the first and second clusters, as indicated by an asterisk (post-hoc test, p = 0.011).

An ANOVA was run using the groups obtained in the cluster analysis and the “caramel – tulip” amygdala activation difference. The results revealed a significant group effect (df=3, F=5.59, p=0.016). Post-hoc comparisons identified a significant difference between the first and the second clusters (p=0.011; Figure 6B). The first cluster was found to have a smaller activation difference for the two odors, and it corresponded to the group with the lowest HQ-CT scores (mean: 3.6) and HQ total scores (mean: 17.4); all of the patients in this group had a HQ-CT score lower than 22, which corresponds to the threshold for clinical severity used in clinical trials. In contrast, patients in the second cluster, who had the largest activation differences for the two odors, all had HQ-CT total scores above 22.

Altogether, our results indicate that there is abnormal, elevated activity in the right amygdala in response to a food-related odor in patients with PWS. This elevated activation relates to hyperphagia and its severity.

## Discussion

This study is the first to investigate odor-related brain activations in adult patients with PWS using fMRI. The results revealed normal performance on an odor recognition test in patients with PWS, but differences in odor-related brain activations. Specifically, we found that for patients with PWS, a food-related odor (caramel) elicited higher levels of brain activation in the right amygdala compared with an odor unrelated to food (tulip); this odor-related difference was significantly higher in the patients than in age-matched controls. The difference between the patient and control groups was large, with the right amygdala activation being five times higher in the PWS group. Three main clusters of patients were identified with respect to the clinical scores of hyperphagia, which correlated highly with the hyperphagia HQ score (r=0.995) and HQ-CT score (r=0.986). These clusters had significantly different right amygdala activations, which indicated that the increased activity related to the degree of hyperphagia. Our results suggest that there may be a specific alteration of the brain circuits involved in processing food odors in PWS, and that this may drive the hyperphagia. It is possible that similar differences may not be apparent for visual processing, which would be in line with our finding that there were no reaction time differences for food and non-food images in patients with PWS, as also found in our previous study (31).

Previous articles have described the concept of “food addiction,” whereby certain foods with added fats or sugars may activate the reward system in a manner similar to drugs of abuse (34-36). This may trigger an addiction to food in susceptible individuals. In line with this, it has been found that high-fat, high-sugar foods are frequently consumed during episodes of binge eating (11, 37, 38), and there is evidence that they may lead to poor control of eating behaviors (37, 39, 40). In addition, foods with added fats and refined carbohydrates are more likely to be intensely craved than are fruit and vegetables (41-43). They are also consumed in greater quantities when people experience negative emotional states (44, 45). Bingeing on these foods leads to changes in the reward system that are seen in other addictive disorders, such as the downregulation of dopamine receptors (46, 47). In our study, caramel was used as the food-related odor, because patients with PWS have a strong preference for sweet foods (33). As this is a high-sugar food, these addiction-related considerations are of particular relevance.

Previous work has shown that odors are potential cues for addictive behaviors (41, 43, 48). Interestingly, the brain circuits involved in processing food odors share many common areas with those activated by cues for addictive substances (49). The key brain areas implicated in food and related emotional/addictive behavior are the basal ganglia, the extended amygdala (central medial amygdala, sublenticular substantia innominata, the nucleus accumbens shell, and the bed nucleus of the stria terminalis), and the prefrontal cortex. In our study, there was hyperactivation in one of these areas, the right amygdala, in response to a sweet food odor in patients with PWS. This finding may relate to the functional connectivity of the amygdala, which may play a role in detecting the emotional salience of different stimuli, particularly those related to food and stress. In line with this, neuroimaging studies in humans have demonstrated increased amygdala activation in response to high-calorie food cues compared with low-calorie food cues (50), and also altered functional connectivity between the amygdala and different cortical areas (51) in obese compared with lean subjects at rest. Furthermore, amygdala activity has been associated with scores on the Yale Food Addiction Scale (27, 28), and both the amygdala and the orbitofrontal cortex have been shown to be more highly activated in obese than lean individuals when anticipating the receipt of food (27). There is evidence that both patients with obesity and those with addictions exhibit augmented activity in the amygdala and ventral striatum in response to reward (52), thus supporting an overlap between food-related and salience-related circuits.

It has been hypothesized that the right amygdala is involved in the rapid detection of emotional stimuli whereas the left amygdala may be more involved in a more sustained evaluation of stimuli (53-55). In support of this, Gläscher et al. found enhanced activation of the right amygdala after the subliminal presentation of emotional facial expressions, while the left amygdala showed increased activation when the stimuli were presented supraliminally (56). In addition, a previous metanalysis found that right amygdala activity was positively associated with craving (57). Our study demonstrated increased activation of the right amygdala in response to a food-related odor in patients with PWS, similar to the increased activation seen in subjects with obesity or substance addiction (52). The right-sided activation is in line with the odors having been presented for a short time and inducing a craving response. We hypothesize that the activations were higher than they would have been for a different food odor with lower sugar content.

Patients with PWS have been found to have high levels of ghrelin and OXT, both of which are involved in addictive behaviors. Interestingly, these hormones have specific regulatory effects on the olfactory bulb in addition to their effects on the hypothalamus and limbic system, including the Ventral Tegmental area (VTA). For instance, OXT has been found to modulate the activity of mitral cells, projection neurons that directly convey sensory information from the olfactory bulb to the olfactory cortices and the medium amygdala (58). In addition, there is increasing evidence that OXT modulates sensory processing, with distinct functions in early and higher olfactory brain regions. The OXT abnormalities in PWS may potentially underlie the increased amygdala activations seen in this study through effects on the olfactory processing pathways. In addition, ghrelin abnormalities may also play a role, as there are ghrelin receptors in the olfactory bulb with neural projections (mitral cells) to the amygdala and hypothalamus. There is evidence that this pathway may be involved in regulating feeding behavior in response to odors (59), and it may be upregulated in PWS.

## Conclusions

Our results show an abnormal hyperactivation of the right amygdala in response to a food-related odor in patients with PWS, which correlates with hyperphagia scores. These results support the notion that the hyperphagia in PWS is an addiction disorder. The findings may relate to abnormalities in food-odor processing that involve the olfactory bulb and dysfunction in the OXT and ghrelin systems.

## Data Availability

All data produced in the present study are available upon reasonable request to the authors

## Acknowledgments and funding

We would like to thank all of the participants in this study, the team at Hendaye Marine Hospital, and the team at the Prader-Willi Syndrome Reference Center at the Children’s Hospital in Toulouse.

This work was supported by grants from the Foundation for Prader Willi Research (ComuFace FPWR OTP 53675) to JD, BP, and MT) and recurrent funding from the Centre National de la Recherche Scientifique (CNRS) (to PB, JD, and KS).

## Author contributions

K.S. - methodology, software, investigation, formal analysis, writing the article, J.D. - methodology, software, investigation, formal analysis, writing the article, J.S. - patient recruitment, formal analysis, writing the article, M.V. - patient recruitment, writing the article,

J.C. - patient recruitment, writing the article, M.T. - conceptualization, methodology, formal analysis, writing the article, P.B. - conceptualization, methodology, formal analysis, writing the article.

## Data sharing statement

Data can be made available upon reasonable request to the corresponding author. The exposure of the data in the online database is not authorized by the hospital.

